# A novel tablet-based software for the acquisition and analysis of gaze and eye movement parameters: a preliminary validation study in Parkinson’s disease

**DOI:** 10.1101/2023.04.04.23288150

**Authors:** Étienne de Villers-Sidani, Patrice Voss, Daniel Guitton, J. Miguel Cisneros-Franco, Simon Ducharme

**Author notes:** **Correspondence** Étienne de Villers-Sidani, 3801 University Rm 742, Montreal, QC, H3A 2B4.

## Abstract

The idea that eye movements can reflect certain aspects of brain function and inform on the presence of neurodegeneration is not a new one. Indeed, a growing body of research has shown that several neurodegenerative disorders, such as Alzheimer’s and Parkinson’s Disease, present characteristic eye movement anomalies and that specific gaze and eye movement parameters correlate with disease severity. The use of detailed eye movement recordings in research and clinical settings, however, has been limited due to the expensive nature and limited scalability of the required equipment. Here we test a novel technology that can track and measure eye movement parameters using the embedded camera of a mobile tablet. We show that using this technology can replicate several well-known findings regarding oculomotor anomalies in Parkinson’s disease, and furthermore show that several parameters significantly correlate with disease severity as assessed with the MDS-UPDRS motor subscale. This tablet-based tool has the potential to accelerate eye movement research via affordable and scalable eye-tracking and aid with the identification of disease status and monitoring of disease progression in clinical settings.

## Introduction

While we have long known that the eyes are our windows to the world, a growing body of research suggests a bi-directional relationship whereby the eyes –and particularly how they move– can also serve as a window into the brain. Although eye movements have previously been linked to certain cognitive processes like attention and decision-making, recent work has unequivocally shown that eye movements can reflect certain aspects of brain function and inform on the presence of neurodegeneration and cognitive impairment (Anderson TJ, MacAskill, 2014; Bueno et al., 2019; Leigh and Zee, 2006; Crotty and Chwalisz, 2019; Terao et al., 2017). The link between eye movements and brain health should not be too surprising given that eye movements are controlled by a diverse network of cortical and subcortical structures (Goffart et al., 2018; Leigh and Zee, 2006) that are susceptible to a variety of degenerative processes (Gorges et al., 2014; Anderson TJ, MacAskill, 2014; Serra et al., 2018). Moreover, the analysis of gaze patterns and visual tasks that measure cognitive inhibition can provide insights into the integrity of various cognitive processes (Liu et al., 2021; Bueno et al., 2019; Fielding et al., 2015).

For instance, two consistent impairments have emerged from Alzheimer’s disease (AD) oculomotor research: a high frequency of saccadic intrusions during attempted fixation and visual capture by the target in the anti-saccade paradigm (Antoniades and Kennard, 2014; Garbutt et al., 2008). Furthermore, microsaccades, tiny horizontal rapid eye movements that interrupt periods of fixation tend to be uniquely obliquely oriented (Kapoula et al., 2014) and occur at an elevated rate in Alzheimer’s (Shakespeare et al., 2015). Parkinson’s Disease (PD) is generally associated with hypometric and multi-step saccades in all types of oculomotor tasks (Terao et al., 2013; Gorges et al., 2016), in addition to a high rate of saccadic intrusions during smooth pursuit (Frei, 2020). Multiple sclerosis (MS) is particularly associated with internuclear ophthalmoparesis (INO)—a slowing of the adducting eye during horizontal saccades—and saccadic intrusions during fixation (Serra et al., 2018). Furthermore, smooth pursuit metrics (low pursuit gain and increased saccadic amplitudes) have been proposed as a marker of early MS (Lizak et al., 2016).

Not only do these oculomotor anomalies characterize these neurodegenerative disorders, but a growing body of research shows that they can serve as markers of disease severity and cognitive impairment. In AD, oculomotor signatures of disease severity have been identified via correlations between specific eye movement characteristics and the Mini-Mental State Examination (MMSE) (Crawford et al., 2005; Noiret et al., 2018; Simó-Servat et al., 2019; Yang et al., 2013). Similarly, in PD, several oculomotor metrics have been shown to correlate with the Unified Parkinson’s Disease Rating Scale (UPDRS) or some of its subscales (Antoniades et al., 2015; Lu et al., 2019; Waldthaler et al., 2019; Zhang et al., 2021), whereas in MS similar relationships have been observed between such metrics and the Expanded Disability Status Scale (EDSS) or the Symbol Digit Modalities Test (SDMT) (Gajamange et al., 2019; Nygaard et al., 2015; Kolbe et al., 2014; Sheehy et al., 2020; Polet et al., 2020).

Although a clinical oculomotor examination is usually sufficient to aid clinicians in the differential diagnosis of advanced neurological disorders, these exams do not typically capture subtle changes such as those highlighted in the aforementioned studies. Indeed, many have proposed that laboratory eye movement recordings can be extremely useful for objective and precise identification of disease status and monitoring of disease progression (Anderson & MacAskill, 2013) and assist with differential diagnoses (Chalkias et al., 2021, Kassevetis et al., 2022; Armstrong, 2015)-though there is hope that the precise quantification of eye movements could also eventually lead to early diagnoses in individuals with less pronounced oculomotor symptoms. Unfortunately, the use of detailed eye movement recordings in clinical settings has been limited due to the expensive nature and limited scalability of the required equipment, such as infrared eye-tracking cameras. While several mobile tablet-based (or smartphone-based) gaze-tracking systems have been developed to provide more accessible and affordable solutions, they, to date, have been limited to the capture and analysis of gross eye movements, such as those required to study gaze search patterns or to determine approximately where an individual is fixating, from which insights regarding memory and attention can be inferred (Haque et al., 2021; Vargas-Cuentas et al., 2017; Bott et al., 2017; Bott et al., 2018).

Eye-Tracking Neurological Assessment (ETNA™) is a recently developed technology that can reliably and accurately track eye movements without the need for infrared cameras, using the iPad Pro embedded camera. This technology allows for the precise quantification of several eye movement parameters currently only available with specialized and costly research-grade infrared eye tracking devices, such as the latency, velocity, and amplitude of saccades, and the presence of saccadic intrusions during fixation. In this paper, we show using the ETNA™ with a standard tablet mobile camera that we can measure and replicate eye movement anomalies and replicate findings from the literature on PD and eye movement, further demonstrating how eye movement parameters can reflect disease status and severity.

## Methods

### Study Design and Subject Population

The Veritas Independent Review Board and the Montreal University Health Center (MUHC) Research Ethics Board gave ethical approval of this work. This cross-sectional study included 121 participants. Fifty-nine (59) PD participants (age 63.76 + 8.23, range 45–79, 35.6 % females) took part in this study. All were recruited by the Quebec Parkinson Network. Inclusion criteria were confirmed diagnosis of PD and sufficient visual acuity to perform the tablet-based visual tasks. Exclusion criteria were the presence of comorbid neurological or psychiatric conditions to avoid eye movement anomaly confounds. To assess clinical status, all PD patients underwent the motor subscale (part III) of the MDS-UPDRS (Goetz et al., 2007; 2008), which was developed to evaluate various aspects of Parkinson’s Disease. Note that because the MDS-UPDRS was performed in a research setting with time constraints and not as part of the standard of care, not all patients underwent the full MDS-UPDRS evaluation. As a result, only part III scores were used in analyses presented herein.

Sixty-two (62) healthy control (HC) participants (age 56.64 + 8.56, range 45–77, 59.7 % females) took part in this study. All were recruited from the Montreal community. The inclusion criterion was sufficient visual acuity to perform the tablet-based visual tasks. Exclusion criteria were evidence or history of other significant neurological or psychiatric disorders. Summary patient demographics are shown in **Table 1**.

**Table 1.** Group demographics.

|  | n | Age | UPDRS part III |
| --- | --- | --- | --- |
| PD patients | 59 | 63.76 ± 8.23 (45–79) | 27.56 ± 13.8 (7–65) |
| Healthy controls | 62 | 56.64 ± 8.56 (45–77) | |

### Gaze-tracking experimental setup

All tests were performed using a 12.9-inch iPad Pro tablet with the ETNA™ software installed, which enables simultaneous video recordings of the eyes and the presentation of visual stimuli on the screen. All participants performed three oculomotor tasks (fixation task, pro-saccade task, and anti-saccade task; see **Figure 1**), which was preceded by a calibration step, where participants were instructed to follow a slowly moving target across the screen. All tablet-based oculomotor tasks were completed in under 6 minutes.

**Figure 1.**
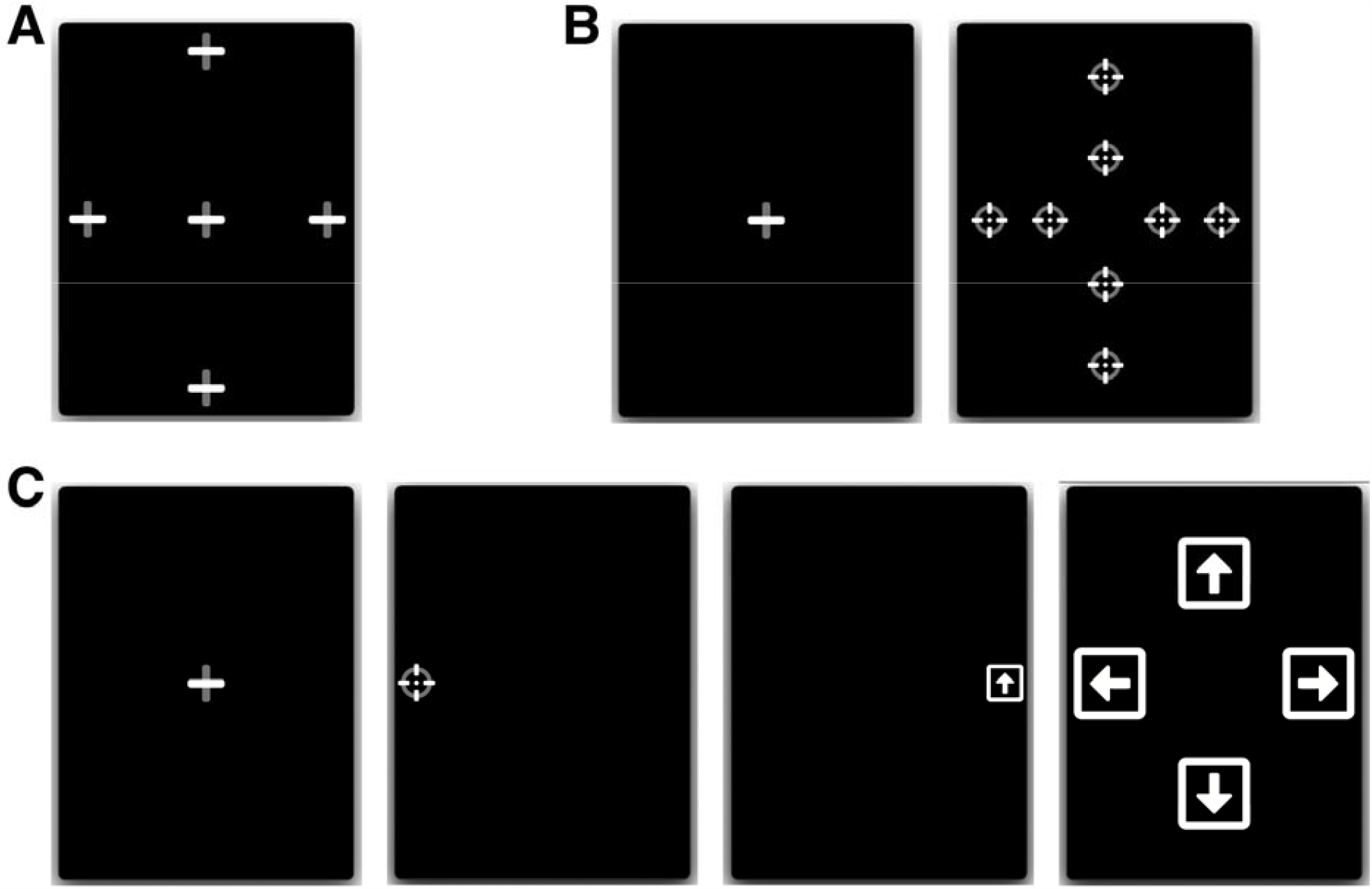
Eye-tracking tasks. (**A**) Fixation: participants fixated a stationary target for 7 seconds, at one of 5 locations. (**B**) Pro-saccades: participants initially fixated a central fixation point, which disappeared after 1.0 – 3.5 s, after which a different target appeared at one of 8 eccentric locations for 1.5 seconds. (**C**) Anti-saccades: participants initially fixated a central fixation point, which disappeared after 1.0 – 3.5 s, after which a round target appeared at 10° to the left or right from the center. Participants were instructed to move their gaze in the opposite direction to the round target, where after 1200ms they were shown a square with an arrow inside that pointed in one of 4 random directions (left, right, up, or down; shown during 400ms). The users then had to direct their gaze towards the arrow orientation corresponding to the arrow they saw in the preceding step.

All tasks were performed with the tablets placed vertically, camera side up, and secured at eye level using a tablet pole mount. Participants were positioned approximately 45 cm from the tablet screen. Safeguards within the gaze-tracking software ensured the participant’s head was properly positioned and visible, at an acceptable angle and distance from the screen.

#### Fixation task

Participants had to fixate a stationary target for 7 seconds, at five different locations (one central and 4 eccentric locations). The eccentric positions were located 10 degrees of visual angle left and right from the center and 14 degrees of visual angle up and down from the center (**Figure 1A**).

#### Pro-saccade task

Participants had to initially fixate a central fixation point, which disappeared after a random period of 1.0 – 3.5 s, after which a different target reappeared at an eccentric location for 1.5 seconds either to the left or right, above, or below the central fixation point. Participants were instructed to move their gaze as quickly as possible to the new target location. Both short (5° horizontal, 6° vertical) and large (10° horizontal,12° vertical) eccentric target distances were used, and each target location was sampled 3 times, for a total of 24 trials (**Figure 1B**).

#### Anti-saccade task

Participants had to initially fixate a central fixation target, which disappeared after a random period of 1.0 – 3.5 s, after which a different target reappeared at an eccentric location (10°) to the left or right from the center. Participants were instructed to move their gaze as quickly as possible in the opposite direction to the new target location. After being displayed for only 100 ms, the target disappeared and the screen was left blank for a predetermined duration of time. Following the blank screen, a symbol appeared in the opposite location of where the initial stimulus appeared (i.e. where the participant should be looking). This symbol consisted of a white square with an arrow inside oriented in one of 4 random directions: either left, right, up, or down. The blank screen period lasted 1200ms and the arrow symbol duration of 400ms. After each trial, a screen was displayed for 5 seconds prompting the user to answer which symbol they saw by directing their gaze toward the arrow orientation corresponding to what they believe is the correct answer (**Figure 1C**). This task was inspired by an anti-saccade task used in a previous study (Guitton et al., 1985), whereby participants could only identify the second symbol had they performed the anti-saccade task correctly (i.e. looked in the opposite direction of the initial target).

### Parameter extraction and analysis

Offline analysis was performed using ETNA™’s proprietary analysis pipeline to automatically extract the eye movement parameters reported for each task. Before parameter extraction, all gaze signals were denoised and non-saccadic artifacts (e.g. blinks) were removed by the software’s analysis pipeline.

The following parameters were extracted from the ***fixation task*** gaze recordings (parameters were averaged across the five fixation trials): 1) 68% bivariate contour ellipse area (BCEA) of fixation - a measure of fixation stability which encompasses an ellipse that covers the 68% of fixation points that are closest to target, 2) 95% BCEA; 3) Horizontal gaze SD - standard deviation of the horizontal gaze position; 4) Vertical gaze SD - standard deviation of the vertical gaze position; 5) the rate of saccadic intrusions during fixation.

The following parameters were extracted from the ***pro-saccade task*** gaze recordings (averaged across all short-eccentricity targets and all large-eccentricity targets): 1) average saccade latency, 2) average total time to reach the target, 3) average mean saccade velocity, 4) average peak saccade velocity, 5) average saccade amplitude gain (amplitude of the saccade relative to the eccentricity of the target; a measure of saccade accuracy), 6) average saccade amplitude error (average distance separating the saccade from the target; a measure of saccade precision)

The following parameters were extracted from the ***anti-saccade task* gaze** recordings: direction error rate, direction corrected rate (proportion of trials where participants directed their gaze in the correct direction following an initial saccade in the wrong direction), target (arrow) recognition rate, correct direction latency, and incorrect direction latency.

Group comparisons were performed using multivariate analysis of variance to simultaneously test statistical differences for multiple response variables (eye-tracking parameters) by one grouping variable (PD or HC). F-statistic with degrees of freedom and p-value are reported. Follow-up between-group comparisons were done using the Kruskal-Wallis test. H-statistic with degrees of freedom and p-value are reported. For correlation analyses with MDS-UPDRS-part III (motor) scores, data normality was assessed with the Shapiro–Wilk test to determine the appropriate correlation coefficient for each eye-movement parameter (i.e., Pearson’s R or Spearman’s ρ). Data analyses and visualization were conducted using R 4.2.1 in RStudio (build 554), packages *dplyr, tidyverse, ggplot2, ggpubr*, and *rstatix*. Although the main purpose of the present paper is to replicate well-known findings in the literature using a novel device, and not to make novel scientific claims, we opted for transparency to present corrected p-values to adjust for the false discovery rate using the Benjamini-Hochberg procedure evaluated at an alpha level of 0.05 (Benjamini & Hochberg, 1995).

## Results

### Group comparisons

Only one significant group difference was observed among the fixation parameters (F(5,121) = 2.64, p = 0.026), where PD patients displayed a higher saccadic intrusion rate (88.7% average increase in PD, H(1) = 19.39, p < 0.001; see also **Table 2** and **Figure 2A**). In contrast, the pro-saccade task yielded several significant group differences (F(14,104) = 14.60, p < 0.001; see **Table 3**), particularly those relating to the number of saccades required to reach the target (14.8% and 23% increase in PD, for short and long eccentricities, respectively; **Figure 2B**), latency (9.7% and 7.2% average decrease in PD, for short and long eccentricities, respectively), saccade precision (13.1% and 9.1% average gain decrease in PD, short and long eccentricities, respectively; **Figure 2C-D**), and mean saccade velocity (20.6% increase in PD, short eccentricity only; **Figure 2E**) (H(1) ≥ 4.69, all p ≤ 0.042).

**Table 2.**
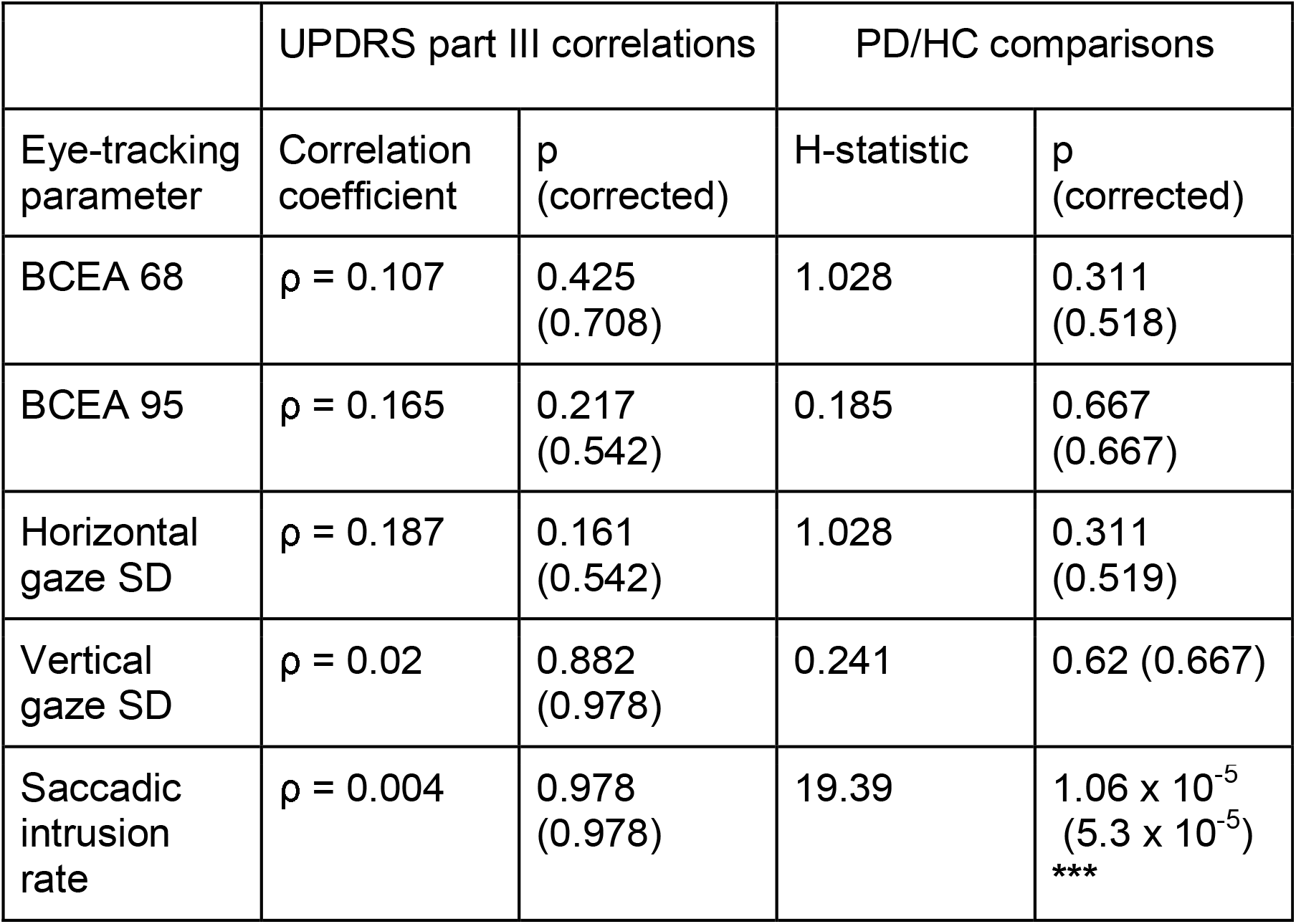
Fixation task. For each eye-tracking parameter, parameter-UPDRS motor score correlations are shown on the left side of the table (*UPDRS part III correlations*). Between-group (PD vs HC) comparisons are shown on the right side of the table (*PD/HC comparisons*). ρ, Spearman’s rho. H-statistic for Kruskal-Wallis test is reported. Raw p-values are presented, followed by their corrected value in parentheses (Benjamini-Hochberg procedure, α = 0.05). *** p < 0.001

**Table 3.** Prosaccade task. For each eye-tracking parameter, parameter-UPDRS motor score correlations are shown on the left side of the table (*UPDRS part III correlations*). Between-group (PD vs HC) comparisons are shown on the right side of the table (*PD/HC comparisons*). ρ, Spearman’s rho. R, Pearson’s R. H-statistic for Kruskal-Wallis test is reported. Raw p-values are presented, followed by their corrected value in parentheses (Benjamini-Hochberg procedure, α = 0.05). * p < 0.05, ** p < 0.01, *** p < 0.001, † p < 0.1

|  | UPDRS part III correlations |  |  |  | PD/HC comparisons |  |  |  |
| --- | --- | --- | --- | --- | --- | --- | --- | --- |
| Eye-tracking parameter | Short amplitude |  | Large amplitude |  | Short amplitude |  | Large amplitude |  |
|  | Correlation coefficient | p (corrected) | Correlation coefficient | p (corrected) | H-statistic | p (corrected) | H-statistic | p (corrected) |
| Latency (mean) | R = 0.263 | 0.048 (0.061) † | R = 0.251 | 0.059 (0.068) † | 17.632 | $2.68 \times 10^{-5}$,<br>($1.25 \times 10^{-4}$) *** | 5.085 | 0.024 (0.038) * |
| Time to target (mean) | R = 0.269 | 0.042 (0.058) † | R = 0.341 | 0.009 (0.02) * | 5.015 | 0.025 (0.038) * | 0.319 | 0.572 (0.604) |
| Saccades to target (mean) | $\rho = 0.153$ | 0.253 (0.253) | $\rho = 0.196$ | 0.142 (0.152) | 41.224 | $1.36 \times 10^{-10}$<br>($1.9 \times 10^{-9}$) *** | 38.313 | $6.02 \times 10^{-10}$<br>($4.21 \times 10^{-9}$) *** |
| First gain (mean) | R = -0.416 | 0.001 (0.007) ** | R = -0.419 | 0.001 (0.007) | 16.099 | $6.01 \times 10^{-5}$<br>($2.1 \times 10^{-4}$) *** | 9.934 | 0.001 (0.002) ** |
| First gain (mean error) | $\rho = 0.371$ | 0.004 (0.014) | $\rho = 0.377$ | 0.003 (0.014) * | 0.919 | 0.338 (0.43) | 7.35 | 0.006 (0.014) * |
| Velocity (mean) | $\rho = -0.339$ | 0.01 (0.02) * | $\rho = -0.334$ | 0.01 (0.02) * | 6.362 | 0.011 (0.022) * | 0.268 | 0.604 (0.604) |
| Peak velocity (mean) | R = -0.271 | 0.04 (0.058) † | R = -0.315 | 0.016 (0.028) * | 4.695 | 0.03 (0.042) * | 0.296 | 0.572 (0.604) |

**Figure 2.**
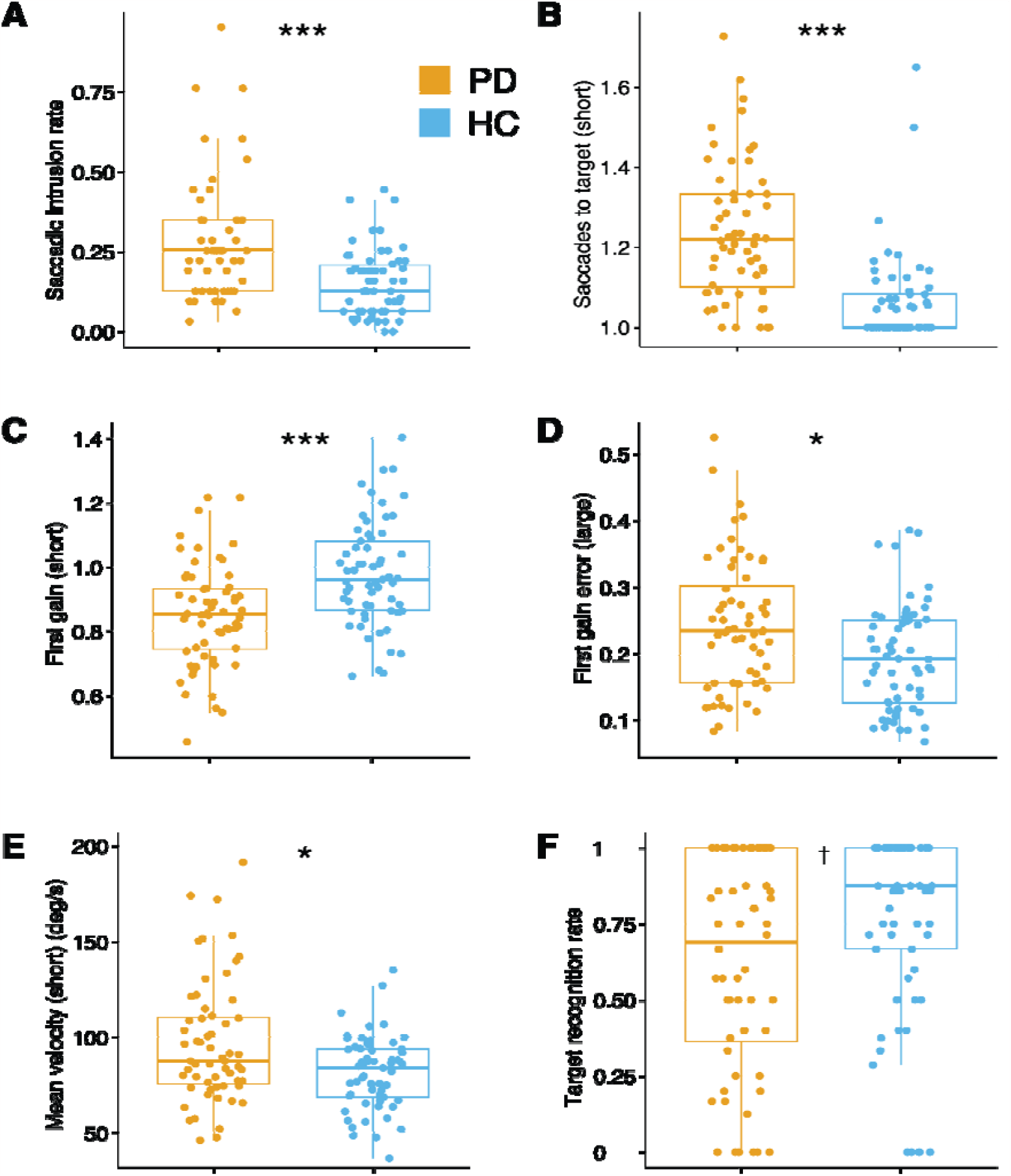
Group differences in eye-tracking parameters between patients with Parkinson’s Disease (PD) and Healthy Controls (HC). Fixation: (**A**) Saccadic intrusion rate; Pro-Saccades: (**B**) saccades to target, (**C**) First gain, (**D**) First gain error, (**E**) Mean velocity; Anti-Saccades: (**F**) Target recognition rate. Large = large amplitude pro-saccades, short = short amplitude pro-saccades. * p < 0.05, *** p < 0.001 (corrected for multiple comparisons) † p = 0.02 (0.10 corrected).

Most parameters for the anti-saccade task did not show any significant group difference, except for the target recognition rate (19.1% decrease in PD; **Table 4** and **Figure 2F**), which implied that although groups did not differ in their proportion of correct direction saccades or in corrected initial direction errors, control participants were on average more frequently correctly identifying the final target.

**Table 4.**
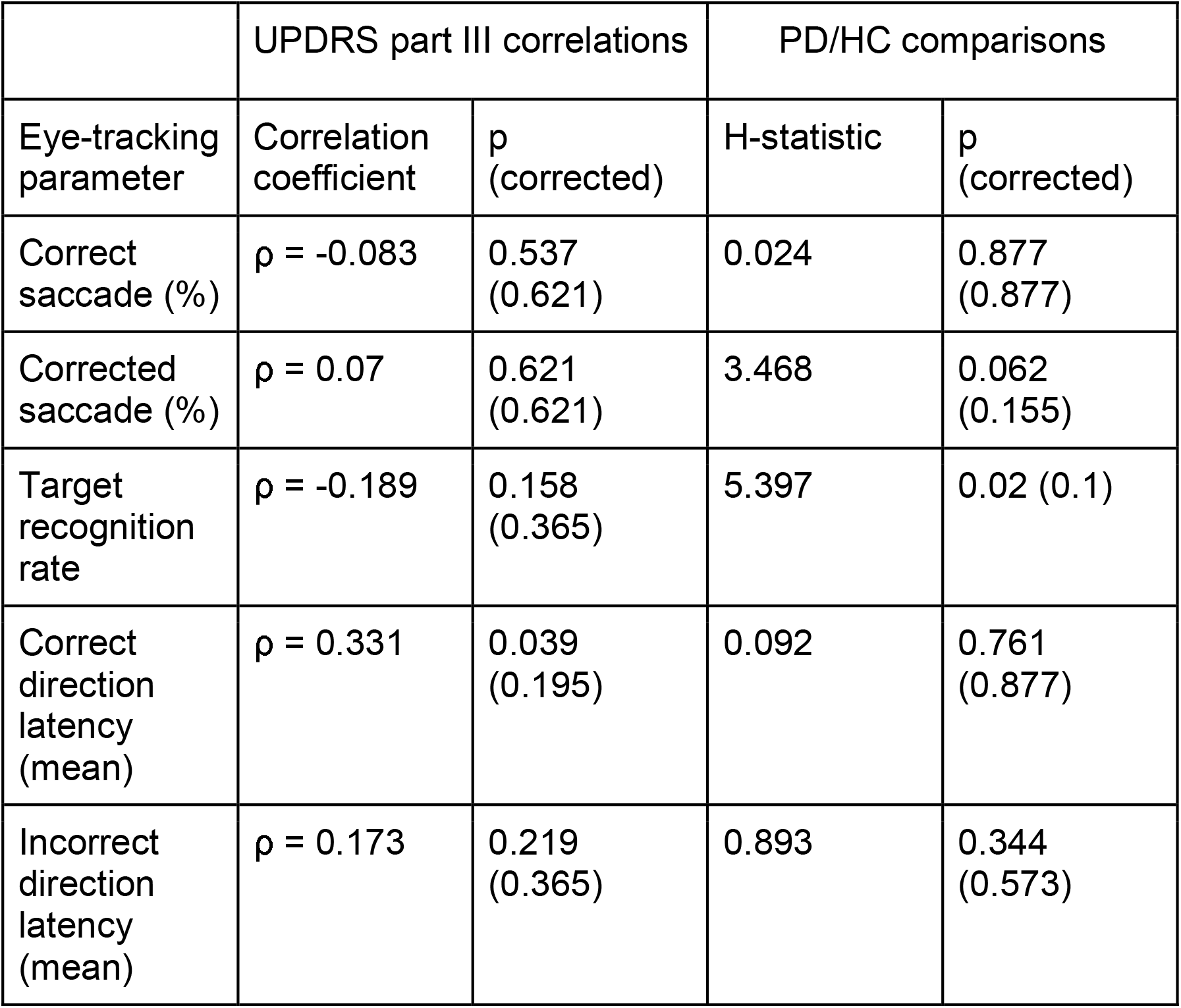
Anti-saccade Task. For each eye-tracking parameter, parameter-UPDRS motor score correlations are shown on the left side of the table (*UPDRS part III correlations*). Between-group (PD vs HC) comparisons are shown on the right side of the table (*PD/HC comparisons*). ρ, Spearman’s rho. H-statistic for Kruskal-Wallis test is reported. Raw p-values are presented, followed by their corrected value in parentheses (Benjamini-Hochberg procedure, α = 0.05).

### Correlations with MDS-UPDRS - part III

No fixation parameters were found to correlate with the UPDRS motor score (all ρ < 0.187, p ≥ 0.542; see **Table 2**). In contrast, most pro-saccade parameters were found to correlate with it, particularly for large eccentricity targets (see **Table 3** and **Figure 3A-E**), six of which survived the p-value correction for multiple comparisons (|R| = 0.315– 0.419, |ρ| = 0.334–377, p ≤ 0.028). A single anti-saccade parameter was found to correlate with the UPDRS motor score (ρ = 0.331, **Figure 3F**), however, the corrected p-value was greater than 0.05.

**Figure 3.**
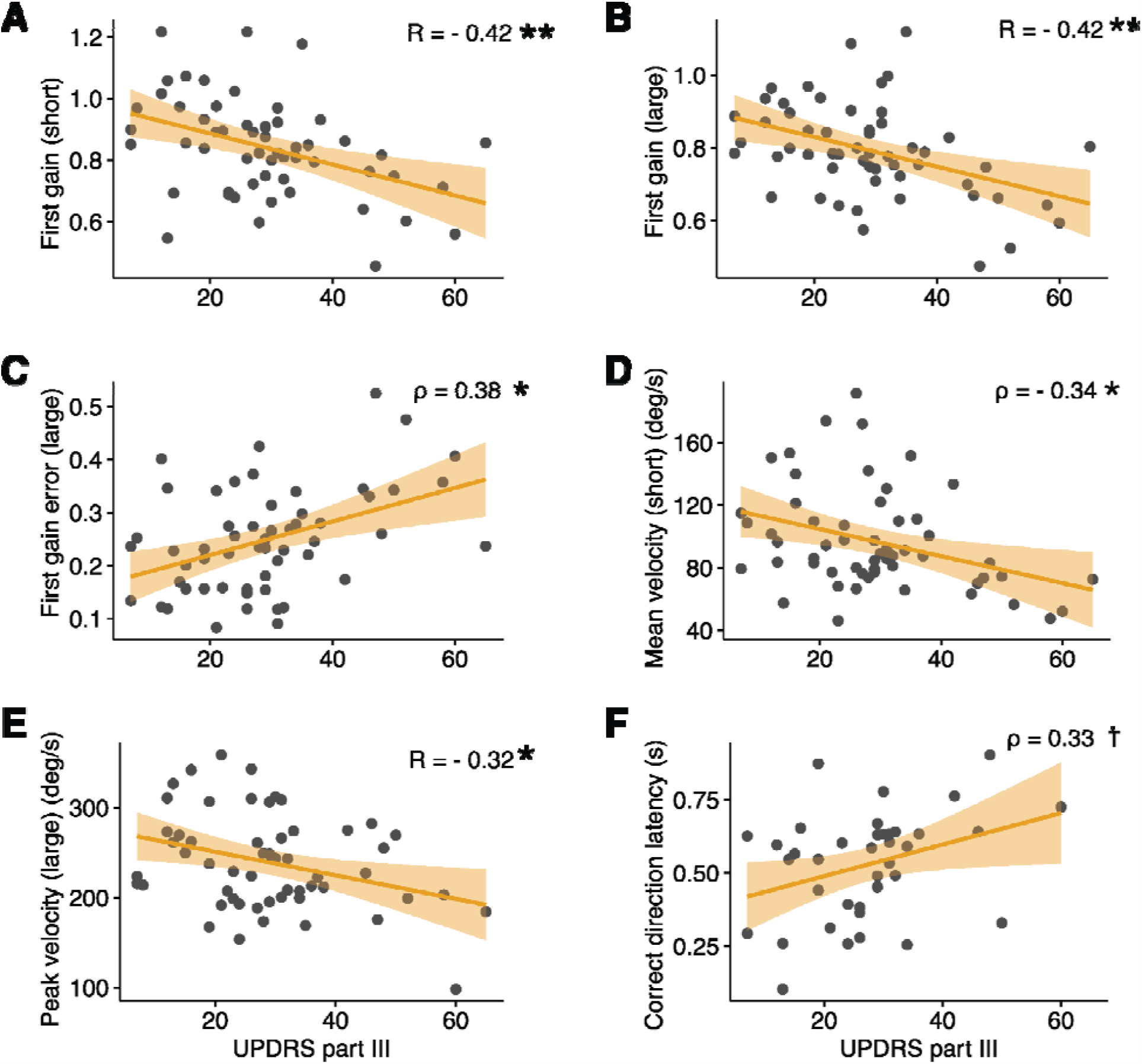
Correlations between select eye-tracking parameters and UPDRS Part III scores. Pro-Saccades: (**A, B**), first gain, (**C**) first gain error, (**D**) mean velocity, (**E**) peak velocity. Anti-Saccades: (**F**) correct direction latency. Large = large amplitude pro-saccades, short = short amplitude pro-saccades. * p < 0.05, ** p < 0.01 (corrected for multiple comparisons) † p = 0.039 (0.19 corrected).

## Discussion

The purpose of the present paper was to demonstrate the potential and usefulness of a novel tablet-based software (currently designed for use with iPad Pros) for the assessment of gaze and eye-movement parameters both in research and clinical practice settings. Indeed, our findings are very much in line with those previously reported in the scientific literature on oculomotor anomalies specific to PD. Our finding of an increased rate of saccadic intrusions during fixation confirms previous reports (Tsitsi et al., 2021; Otero et al., 2013; Altiparnak et al., 2006; Zhang et al., 2021). With regards to measures of gaze stability, to our knowledge, only one study reported BCEA and horizontal/vertical gaze SD measures in PD patients and found no significant differences with healthy controls (Tsitsi et al., 2021).

Pro-saccades have been more extensively studied in PD than measures of fixation instability. Findings regarding saccade latency have been mixed to date, with several studies finding either no differences between PD patients and controls (Gitchel et al., 2012; Blekher et al., 2009, Perkins et al., 2021; Nagai et al., 2020), shorter latencies in PD (Zhou et al., 2021) and longer latencies in PD (Zhang et al., 2021; Wu et al. 2020; Waldthaler et al., 2019). It’s unclear why this discrepancy across studies exists, but it may have to do with the type of eye-tracking technology used; the majority of the studies cited above that found either no difference or shorter latencies in PD used infrared eye-trackers from manufacturers such as Eyelink and Tobii, whereas the majority of studies cited above finding increased latencies used devices from other manufacturers such as Micromedical Technologies and Interacoustics, and EyeBrain. To make a more definite statement as to why this would be the case, however, would require a more in-depth investigation that is beyond the scope of the paper.

Similarly, findings regarding peak velocity have been mixed, with a few studies finding faster velocities in PD (Lu et al., 2019; Grillini et al., 2020), whereas most other studies found no differences between PD and healthy control (Blekker et al., 2009; Stuart et al., 2013; Gorges et al. 2013; Zhou et al., 2021; Gitchel et al., 2012). In the present study, we find peak and mean velocities to be significantly increased in PD patients (only uncorrected p-value for peak velocity) for short eccentricity targets only. Combined with the available literature, this finding suggests that peak velocity might normalize in PD with increasing eccentricity.

In contrast to latency and velocity parameters, the literature is quite rich with strong evidence of PD patients requiring multi-step saccades to reach the target (Shaikh et al., 2019, Blekher et al., 2009, Zhou et al., 2021 Gorges et al. 2013), which is in line with the present findings reported here, where the average number of saccades required to reach the pro-saccade targets was significantly greater in PD patients. In addition, our findings here indicate that the first saccade towards the target (for both short and large eccentricity targets) was closer to the target in HC.

With regards to anti-saccade parameters, although several studies report a reduced proportion of correct initial direction (or an increase in error rate) in PD patients (Nagai et al., 2020, Terao et al., 2019; Waldthaler et al., 2019; Barbaso et al., 2019; Ouerfelli-Ethier et al., 2018; Gorges et al., 2017), several other studies found no such difference (Pagonabarraga et al., 2021; Ewenczyk et al., 2017; Visser et al. 2019). Aligned with this discrepancy, our findings landed somewhat in the middle, with the group difference lying just above the statistical significance threshold. Similarly, while several studies have identified slower latencies for correct (Nagai et al., 2020, Terao et al., 2019; Ewenczyk et al., 2017), others found no group differences (Ouerfelli-Ethier et al., 2018; Gorges et al., 2017; Barbaso et al., 2019).

Taking a closer look at the reported findings in the literature, it can be observed that many of those studies that identified a difference in the correction direction rate found no differences regarding latency (Ouerfelli-Ethier et al., 2018; Gorges et al., 2017; Barbaso et al., 2019), and vice-versa (Waldthaler et al., 2019; Ewenczyk et al., 2017; Visser et al. 2019), indicating either potentially large variability in the PD population or that the differences measured could be specific to the anti-saccadic task parameters

(e.g. eccentricity of the targets or inter-trial interval). A recent meta-analysis on antisaccade parameters in PD confirmed that, although both antisaccade latency and error rate are significantly increased in PD, these effects are strongly moderated by disease duration and, as assessed by UPDRS score and H&Y stages, disease severity, (Waldthaler et al., 2021). This likely explains the absence of significant findings regarding the antisaccade latency and error rate in the present study, as the majority of our PD participants would fall in the mild or moderate category based on their MDS-UPDRS score part III (Martínez-Martín et al., 2015). However, we did find a significant group difference (uncorrected p-value only) regarding the ability to correctly identify the second target (arrow symbol), and the group difference for the *percent corrected* rate (i.e. the rate at which participants executed a saccade in the correct direction after an initial saccade in the incorrect direction) was just above above the statistical significance threshold. These two more subtle measures of impairment that are typically not reported in the literature, and suggest that HC are more likely to correct their initial anti-saccade error to detect and recognize the second target.

Few studies to date, to our knowledge, have investigated the relationship between disease severity in PD, such as measured by the MDS-UPDRS motor score and the magnitude of gaze and eye movement parameters. These have primarily observed a relationship between the motor score and pro-saccade latency (Zhang et al., 2021; Zhang et al., 2018), prosaccade gain (Waldthaler et al., 2019), anti-saccade latency (Lu et al., 2019) and anti-saccade direction rate (Antoniades et al., 2015). However, Visser et al., (2019) found no significant correlation between anti-saccade latency or pro-saccade latency and the UPDRS motor score.

In the present study, we only found a significant UPDRS motor score correlation with pro-saccade gain (large eccentricities) and the number of saccades to reach the target (large eccentricities). We also found a significant correlation between the UPDRS motor score and the pro-saccade time-to-target parameter (large eccentricities), which in many ways represents a composite measure of the latency and the mean velocity of the saccade. With regards to the anti-saccade task parameters, it is unclear why the discrepancies between the cited literature and our study exist. One obvious difference between our PD patient sample is that the error rate was significantly larger in our study (61% vs 15% in Antoniades et al., 2015) despite anti-saccade targets being positioned at a similar eccentricity.

Despite the promise of eye tracking for both research and clinical settings, applications have been limited by the high cost of eye trackers and their inability to scale due to the use of specialized hardware. Being able to make use of the embedded cameras of mobile devices allows us to overcome these cost and scalability barriers by democratizing access to eye-tracking assessment tools. In particular, we believe tablet-based tools have the potential to aid with disease progression monitoring via the assessment of the integrity of the oculomotor system, as demonstrated by the strong relationships found between various eye-movement parameters and clinical status. Such tools could help clinicians monitor changes to disease status, disease progress, or response to treatment remotely without the need for an in-clinic visit until a change in associated eye movement parameters is detected by the software. This approach would be akin to current alternative strategies being developed to remotely monitor motor function & dysfunction with gyroscope/accelerometer-based wearable technologies (Rodríguez-Molinero et al., 2018; Tripoliti et al., 2013) and speech analysis using machine learning techniques (Almeida et al., 2019; Rahman et al., 2021).

An advantage of eye-movement-based monitoring technologies, as opposed to wearable technologies, for example, is that they could potentially be more easily scaled to other neurodegenerative disorders. Indeed, as highlighted earlier, several eye-movement anomalies have been tied to AD (Shakespeare et al., 2015, Antoniades and Kennard, 2014) and MS (Serra et al., 2018; Lizak et al., 2016), for instance, and several measured parameters have been shown to highly correlate with their respective cognitive (Noiret et al., 2018; Simó-Servat et al., 2019; Yang et al., 2013) or clinical disease scales (Gajamange et al., 2019; Nygaard et al., 2015; Kolbe et al., 2014; Sheehy et al., 2020; Polet et al., 2020).

To conclude, in this study we show that a novel tablet-based eye-tracking technology can reliably identify differences in subtle eye movement abnormalities in PD, and that specific oculomotor parameters were found to significantly correlate with the disease severity stage. Next steps include validating the technology within a distinct neurodegenerative disorder with known oculomotor impairments. This tablet-based tool has the potential to rapidly scale eye-tracking use and usefulness in both research and clinical settings.

## Data Availability

All data produced in the present study are available upon reasonable request to the authors

## Acknowledgements

We would like to express our sincere gratitude to Edward Fon, Clotilde Degroot, and Roozbeh Sattari from the Quebec Parkinson Network for their valuable contributions to this research project. We also thank Jimmy Lai, Redouane Allache, Sarah Fon, Lydia Ouellet, Fama Tounkara, and Thedora Yaneva for their help with data acquisition.

## Contributions

CRediT (Contributor Roles Taxonomy) author statement: **Conceptualization**: E.d.V.-S., S.D.; **Formal analysis**: P.V., J.M.C.-F.; **Funding acquisition**: E.d.V.-S.; **Investigation**: J.M.C.-F., S.D.; **Methodology**: E.d.V.-S., P.V., D.G.; **Supervision**: E.d.V.-S., S.D.; **Visualization**: J.M.C.-F.; **Writing – original draft preparation**: P.V.; **Writing – review and editing**: E.d.V.-S., P.V., D.G., J.M.C.-F., S.D.

